# The Effects of Social Distancing Policy on the Changes of Floating Population in Korea

**DOI:** 10.1101/2024.08.26.24312613

**Authors:** Jaehwan Oh, Minsu Choi, Kwang-soo Lee

**Author notes:** **Corresponding author(s):** Kwang-soo Lee, Department of Health Administration, Yonsei University Graduate School, Changjo Hall, Room Number 413, Yonseidaegil 1, Wonju-si, Gangwon-do, Republic of Korea. This paper is a partial revision of the researcher’s master’s thesis (Jaehwan Oh, Yonsei University Graduate School, 2023).

## Abstract

**Objectives:** In response to the World Health Organization’s declaration of the COVID-19 pandemic in March 2020, nations worldwide, including Korea, implemented social distancing as a critical Non-Pharmaceutical Intervention (NPI) to curb the spread of the disease. Social distancing measures aimed to reduce person-to-person contact through various strategies such as facility restrictions, gathering limitations, travel bans, and lockdowns.

**Methods:** This study investigated the impact of social distancing policies on the floating population across 229 administrative districts in Korea. The dependent variable was the total floating population, while the independent variable was the social distancing stage of the week, focusing on stages that prevailed for more than half of each week. Control variables included sex ratio, season, and the number of weekly holidays.

**Results:** Descriptive analysis and t-tests were conducted to examine the variables, and Panel GEE analysis was performed to analyze changes in the floating population with the social distancing stage. The analysis revealed a significant decrease in the floating population when transitioning from stage 5 to stage 4 of social distancing. This indicates that stricter gathering restrictions and increased local government autonomy effectively reduced person-to-person contact.

**Discussion:** These findings underscore the importance of targeted social distancing measures in mitigating transmission risks during infectious disease outbreaks. The study provides valuable insights for future policymaking on infectious diseases, offering relevant data to inform effective public health strategies and responses. Understanding the impacts of such measures can help refine approaches to managing future pandemics and ensuring public health safety.

## Introduction

Coronavirus Disease (COVID-19) spread worldwide rapidly with high contagiousness. World Health Organization (WHO) declared it a pandemic in March 2020.

In January 2020, Korea confirmed its first COVID-19 case and the Korea Center for Disease Control and Prevention (KCDC) traced and isolated infected individuals (1). However a massive infection in the Daegu church community in February 2020 led to a nationwide spread, making Korea’s cumulative cases second only to China^2^. This high contagiousness necessitated new treatments and protective measures, including social distancing policies.

According to Kim (2), this policy was implemented to reduce contact between individuals. Characteristics with close contact increase the probability of infection. Therefore, to alleviate the spread of disease in local communities, it must avoid contact with individuals in a close environment as much as possible. The pattern of infection has changed due to the continued COVID-19, and social distancing stage 4 has been applied since July 1, 2021, to construct a sustainable social distancing system(3). Previous studies have shown the effectiveness of social distancing in reducing COVID-19 spread, often measured by changes in population movement(4, 5). However, these studies were limited in scope and duration.

Therefore, this study aimed to analyze whether the social distancing policy significantly affected the floating population in 229 administrative districts in Korea as the social distancing changed from stage 5 to stage 4.

### Concept of Social Distancing

The social distancing policy is one of the NPIs(Non-Pharmaceutical Interventions), that considers the characteristics of infectious diseases infected through contact between people. It aims to reduce person-to-person contact through restrictions on the use of facilities, limiting the number of people gathering, travel bans, and lockdowns to prevent the spread of disease. Maragakis (6) reported that social distancing means staying home and away from others as much as possible to help prevent the spread of COVID-19. The Centers for Disease Control and Prevention (CDC) has described social distancing as a method for reducing the frequency and closeness of contact between people to decrease the risk of disease transmission (7).

### Specific information on social distancing in Korea

Korea’s initial sporadic social distancing policies were reorganized into three stages by June 2020(8). By November 2020, stage 5 was introduced, subdivided into five levels to address prolonged COVID-19 threats(9). Stage 4, implemented nationwide from July 2021, gave local governments the autonomy to adjust measures based on regional circumstances(3).

## Methods

### Study framework and data

In this study, the researcher investigated the impact of social distancing changes on the number of floating populations in regions. As shown in <Figure 1>, the dependent variable was the number of floating populations, while the independent variable was the changes in social distancing. Control variables, including socio-demographic factors, seasonal factors, and the number of holidays per week, were considered due to their potential influence on the floating population. The model aimed to assess whether transitions in social distancing stages from 5 to 4 had significant effects on the number of floating populations across regions.

**Figure 1.**
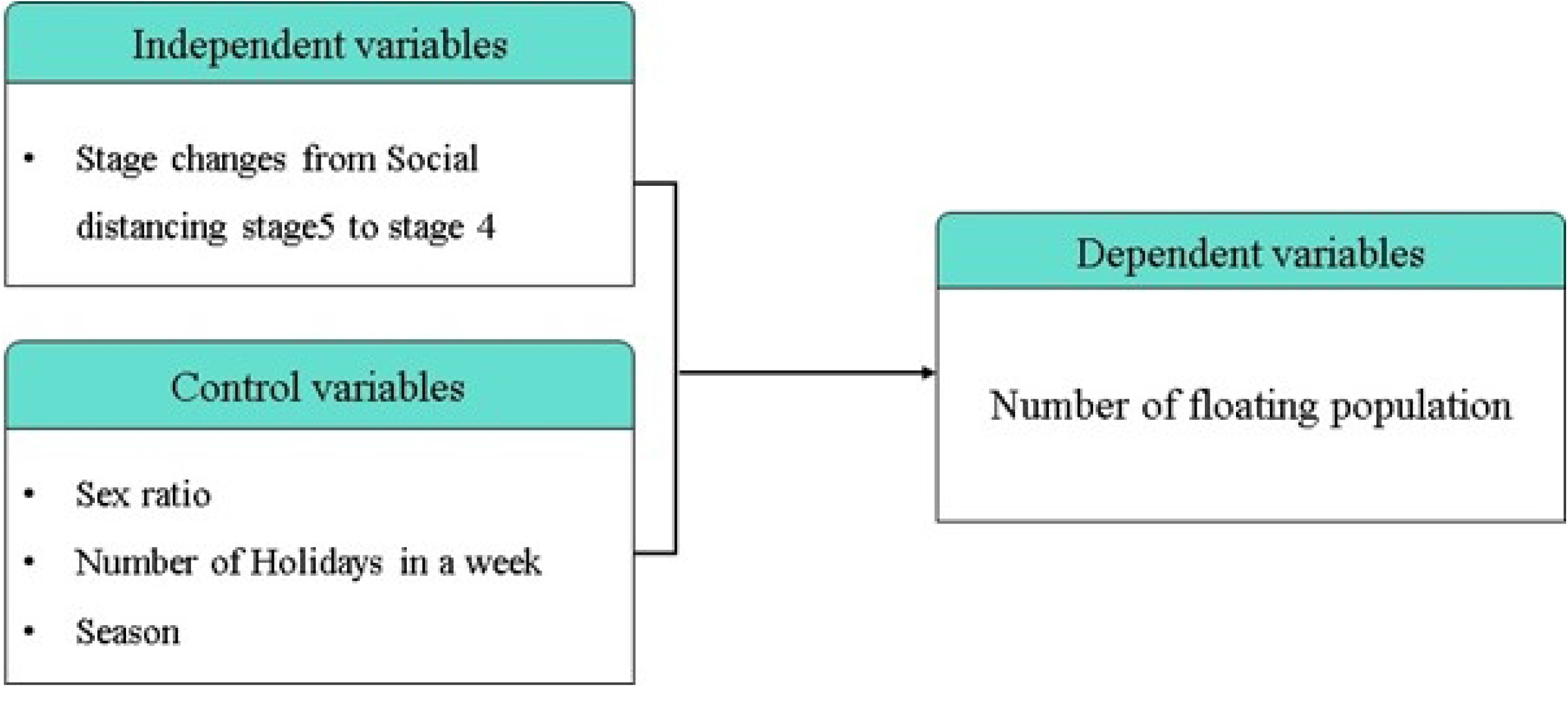
Study framework

### Data

The dependent variable is the total number of floating population in 229 administrative regions in Korea. In contrast, the independent variable is the social distancing stage changes from stage 5 to 4. Control variables include sex ratio, season, and number of holidays in a week.

Floating population data were sourced from mobile statistics provided by Statistics Korea and SK Telecom. The population was counted weekly by region, with movements recorded if a mobile signal moved to another area and stayed for over 30 minutes. Jang (10) noted that the average daily number of movements was also calculated. The total floating population included both incoming and outgoing movements, estimated by SK Telecom using demographic ratios.

Social distancing data were obtained daily from the Ministry of Health and Welfare via Open API. Weekly stages were determined by the predominant stage for more than half of the week. For instance, if stage 4 was applied for most of the week, it was recorded as such.

Control variables were based on previous studies. The sex ratio, indicating the number of men per 100 women, was obtained from the Korean Statistical Information Service (KOSIS). Seasonal effects were categorized as spring (March-May), summer (June-August), autumn (September-November), and winter (December-February) (11). The number of holidays per week was calculated by summing national holidays, Saturdays, and Sundays(12). Regional data were sourced from the 2014 census boundary data provided by the Statistical Geographic Information System (SGIS).

### Statistical Analysis

Descriptive analysis and t-test were conducted to examine the variables. Panel GEE(Generalized Estimating Equation) was conducted to analyze floating population changes with the social distancing stage. STATA 15.0 (S. E) was used for statistical analysis.

A Generalized Estimating Equations (GEE) model accounts for autocorrelation within a dataset with repeatedly measured observations. According to Diggle (13), GEE enables the examination of the correlation among dependent variables across time points. As per Park and Jeon (14), notably, the panel GEE model operates without imposing strict assumptions on parameter distributions and can be applied even in cases where the distribution of the dependent variable is unknown or violated. GEE produces asymptotic estimates leveraging quasi-likelihood functions, utilizing information solely from the distribution and correlation of response variables. Per Zorn (15), by employing Generalized Linear Models (GLM), GEE accommodates variables that deviate from normal distribution and facilitates the consideration of autocorrelation within the dataset.

## Results

### General statistics for study variables

The trend of the average floating population during the study period is shown in <Figure 2>. There were no consistent trends in the floating population; however, it decreased continuously from the 30th week when stage 4 of social distancing was applied nationwide.

**Figure 2.**
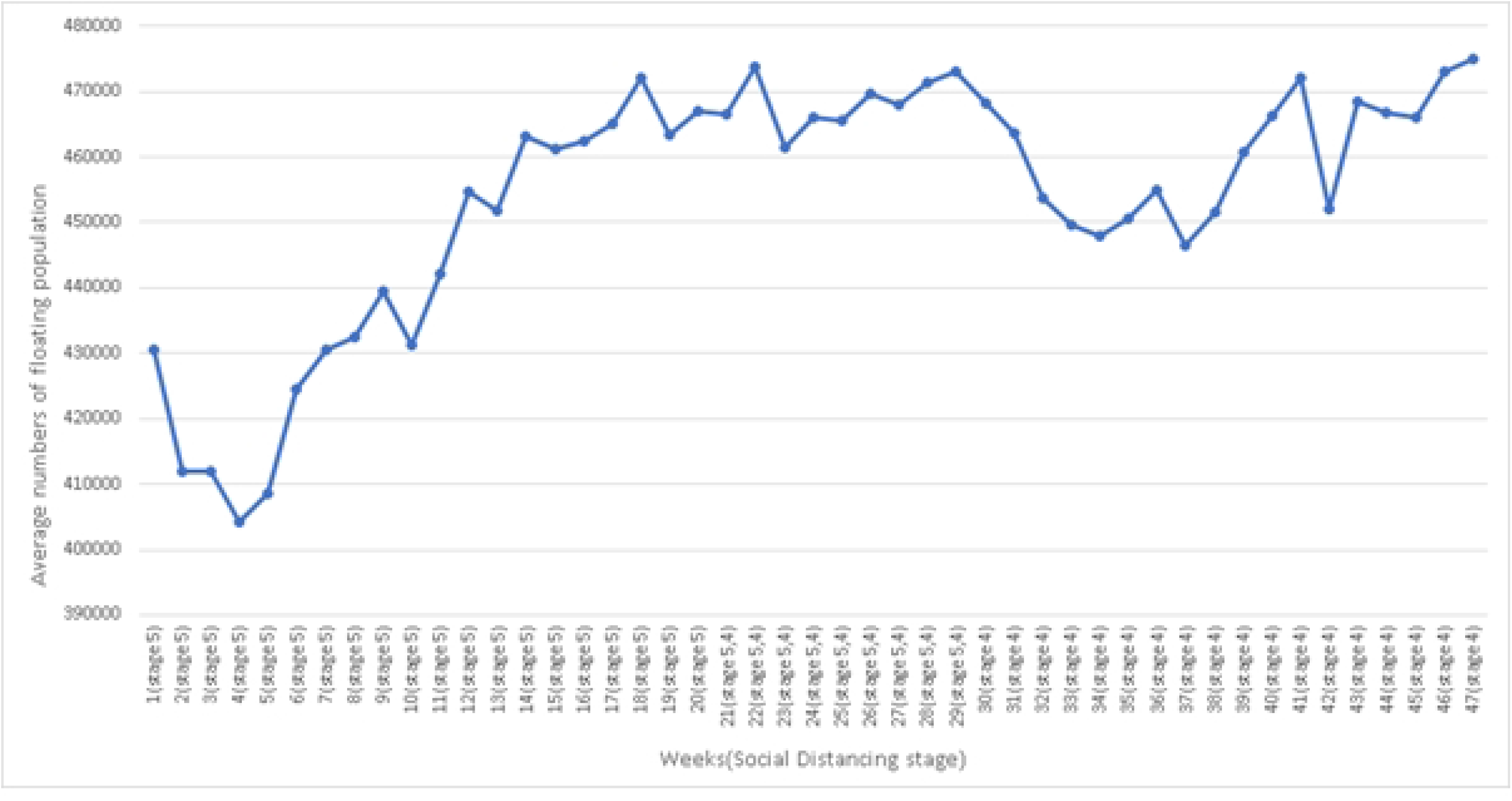
Trend of the average floating population nationwide by week

### Difference analysis

T-test was conducted to analyze, whether there were significant differences in study variables between stages 4 and 5 of social distancing. Table 1 shows significant differences in the number of holidays and the number of floating population between the two periods.

**Table 1.** Differences of variables between stage 4 and 5.

|  |  |  | [Mean (SD)] |
| --- | --- | --- | --- |
| Variables | Stage |  | t-value |
|  | Stage 5 | Stage 4 |  |
|  | (N=6,255)<sup>1)</sup> | (N=4,508)<sup>2)</sup> |  |
| Number of Floating | 464304.41 | 428010.19 | 4.34*** |
| Population | (432228.70) | (421638.05) |  |

\* p<.05, \*\* p<.01, \*\*\* p<.001
1) The number of regions where the social distancing stage 5 was applied during the study period.
2) The number of regions where the social distancing stage 4 was applied during the study period.

### Panel GEE

Table 2 presents the results of Panel GEE analysis examining the effects of transitioning from social distancing stage 5 to stage 4. Notably, there was a significant average decrease of 3,884.52 in the floating population following this shift in social distancing measures. Seasonal variations were evident, with increases observed during spring, summer, and autumn compared to winter.

**Table 2.** Results of Panel GEE Analysis.

| Variables | Coefficient | z-value |
| --- | --- | --- |
| Social Distancing (Ref: stage 5) |  |  |
| Stage 4 | -3,884.52 | -3.58*** |
| Season (Ref: winter, December to February) |  |  |
| Spring (March to May) | 17,117.74 | 16.06*** |
| Summer (June to August) | 23,501.41 | 17.29*** |
| Autumn (September to October) | 34,291.33 | 21.63*** |
| Sex ratio | -6,223.60 | -2.48** |
| Number of holidays in a week | -4,218.14 | -19.23*** |

\* p<.05, \*\* p<.01, \*\*\* p<.001

Furthermore, the analysis revealed significant associations. The sex ratio demonstrated a noteworthy impact, with an average decrease of 6,417.94 observed in the floating population with each unit increase in the sex ratio. Similarly, an increase of 1 unit in the number of holidays corresponded to an average decrease of 4,218.14 in the floating population.

These findings underscore the influence of social distancing policies, seasonal trends, and demographic factors on population mobility dynamics during the specified period.

## Discussion

In Korea’s 229 Si. Gun. Gu regions, the floating population did not exhibit specific patterns and decreased during the transition from social distancing stage 5 to stage 4 in the 30th week. Additionally, the number of regions under stage 4 of social distancing varied sporadically during the initial phase of the study period. These changes occurred between week 30 to week 34 and remained consistent until the conclusion of the study period, from week 35 onwards.

On the other hand, all variables used in Panel GEE analysis significantly affected the number of floating population. The possible explanation is as follows.

The Ministry of Health and Welfare (3) has proposed reforms to social distancing protocols in response to increased infections linked to contact with confirmed cases. Consequently, rigorous measures were implemented during social distancing stage 4, including restrictions on private gatherings, face-to-face religious gatherings, rallies, and the size of gatherings. Moreover, during stage 4-4, the number of individuals allowed at private gatherings was further limited to 2 people after 6 p.m. These heightened activity restrictions likely played a role in reducing the floating population.

Flexible social distancing policies, which take into account regional epidemic levels and local government capabilities, may have contributed to the reduction in the floating population. According to the Ministry of Health and Welfare (3) During social distancing stage 5, the central government dictated the social distancing measures. In contrast, in stage 4, local governments had the authority to apply social distancing flexibly based on regional circumstances. For instance, on July 1st, 2021, Gangwon-do was at stage 4-1, while Chuncheon-si in Gangwon-do implemented stage 4-3. Similarly, during the first week of July, Namhae-gun and Tongyeong-si upgraded their social distancing stage to 4-2 due to a continuous increase in confirmed cases, even though Gyeongsangnam-do, including Namhae-gun and Tongyeong-si, remained at stage 4-1. In early October 2021, Gyeongsangbuk-do was at stage 4-3, but 12 regions, including Uljin-gun, adopted stage 4-1. However, Uljin-gun enforced stricter standards for private gatherings, following the trend of rising COVID-19 cases in other regions (16).

Effective disease prevention and control policies, such as lockdowns, were implemented by local governments in New York(17). No secondary severe outbreaks were observed until November of the same year. Previous studies have also suggested that the direction of COVID-19 spread in large cities varies depending on the response policy or method. Gerritse (18) stated that while a government-centered COVID-19 response method may have advantages, considering the information and resources of local governments, it is suggested that local governments have an appropriate level of autonomy in responding to infectious diseases.

Seasonal variations significantly influence the mobility of individuals. A previous study demonstrated that walking activities tend to increase during spring and summer compared to winter (19). As stated by Shin (20), the implementation of the “Support Plan for Daily Recovery of Vaccinated People” on May 26th is anticipated to augment the floating population. Individuals who have received their first vaccination are exempted from limitations on the number of attendees for direct family gatherings and religious activities. Additionally, they enjoy benefits such as the removal of masks during outdoor activities, likely leading to an increase in the floating population during the summer months. Conversely, Snoeijer, Burger (21) have indicated that non-pharmaceutical interventions (NPIs) like lockdowns decrease mobility. Therefore, easing such interventions could potentially contribute to an increase in the floating population.

The results revealed a significant decrease in the floating population as the sex ratio increased. Kim, Lee (22) have suggested that women constitute a higher percentage of the floating population in densely populated areas due to a smaller share of women workers compared to men. According to Statistics Korea (23), in the Statistics Korea’s Economic Activist Survey, there were 16,124,000 economically active men and 12,186,000 women in 2021. Furthermore, the Ministry of Employment and Labor (24) announced that the number of telecommuters surged from 95,000 in 2019 to 1.14 million in 2021 due to the COVID-19 pandemic. These structural shifts may have led to an increase in male telecommuting, which could also impact the floating population dynamics.

## Conclusion

This study examined the impacts of social distancing on the floating population across 229 regions of Korea from December 7, 2020, to October 31, 2021.

The floating population experienced a significant decline when social distancing transitioned from stage 5 to stage 4. Compared to winter, the floating population increased in all seasons. Moreover, notable changes were observed in the sex ratio and season during this period. The variance in the floating population between stage 4 and stage 5 of social distancing can be attributed to stricter restrictions and increased autonomy granted to local governments. The policy regarding gatherings was reinforced in stage 4 compared to stage 5, suggesting that maintaining regulations could lead to a decrease in the floating population. Ko, W. (17) have indicated instances where local governments effectively prevented and controlled disease spread using their capabilities. Hence, local governments should be afforded a certain level of autonomy to implement disease control and prevention measures tailored to regional circumstances.

## Data Availability

The data underlying the results presented in the study are available from (Floating population: https://data.kostat.go.kr/social/moblilePopMoveInfoPage.do) (Social Distancing: https://www.data.go.kr/data/15098772/openapi.do?recommendDataYn=Y (Open API)) (Sex ratio: https://kosis.kr/statHtml/statHtml.do?orgId=101&tblId=DT_1B040A3) (Regional Data: https://sgis.kostat.go.kr/view/pss/openDataIntrcn#).

https://kosis.kr/statHtml/statHtml.do?orgId=101&tblId=DT_1B040A3

https://www.data.go.kr/data/15098772/openapi.do?recommendDataYn=Y

https://data.kostat.go.kr/social/moblilePopMoveInfoPage.do

## References

1. Hyun J-H, Kim J-H, Lee HY, Gwack J, Kim J-E, Lee E-Y, et al. Contact Tracing Results of the First Confirmed COVID-19 Case in the Republic of Korea. Public Health Weekly Report. 2020;13(7):352∼8.

2. Kim NS. Current status and challenges of COVID-19. Ministry of health and welfare Issue & Focus. 2020;373:1–13.

3. Reorganization of the Social Distancing System (Proposal) [press release]. Korea Policy Brefing, June 2021.

4. Koo JR, Cook AR, Park M, Sun Y, Sun H, Lim JT, et al. Interventions to mitigate early spread of SARS-CoV-2 in Singapore: a modelling study. Lancet Infect Dis. 2020;20(6):678–88.

5. VoPham T, Weaver MD, Hart JE, Ton M, White E, Newcomb PA. Effect of social distancing on COVID-19 incidence and mortality in the US. medRxiv. 2020.

6. Maragakis LL, M.D., M.P.H. Coronavirus, Social and Physical Distancing and Self-Quarantine Johns Hopkins Medicine2020 [updated 16, Feb. 2022. Available from: https://www.hopkinsmedicine.org/health/conditions-and-diseases/coronavirus/coronavirus-social-distancing-and-self-quarantine.

7. Kinlaw K, Levine R. Ethical guidelines in pandemic influenza. Centers for Disease Control and Prevention. 2007.

8. Kim CK. The government classifies and implements three stages of ‘social distancing’… Transition according to the degree of spread. Korea Policy Briefing. 2020.

9. Central Disaster and Safety meeting [press release]. Korea Policy Briefing: Office for government policy coordination, November 2020.

10. Jang YH. Reading the characteristics in terms of traffic volume by purpose 1, Centered on the statistical data of the SDC telecommunication mobile population movement of the Statistics Korea. Planning and Policy. 2022;488:72–7.

11. Lee SM, Hong SJ. The Effect of Weather and Season on Pedestrian Volume in Urban Space. Journal of Korea Academia-Industrial Cooperation Society. 2019;20(9):55–65.

12. Son S, Seo C. Chuseok Holiday Experiences and Prospects of Changes in the Culture of Holidays after the COVID-19 Pandemic. Society for the Study of Family Policy. 2021;1(2):55–74.

13. Diggle PJ. Introduction to Liang and Zeger (1986) Longitudinal Data Analysis Using Generalized Linear Models. In Breakthroughs in Statistics. Springer. 1997:463–82.

14. Park CS, Jeon YM. Bootstrap Estimation for GEE Models. Korean Journal of Applied Statistics. 2011;24(1):207–16.

15. Zorn CJW. Generalized estimating equation models for correlated data_A review with applications. American Journal of Political Science. 2001;45(2):470–90.

16. Joo HS. Uljin-gun, distancing level 1 and ban on private gatherings of 5 or more people extended by 2 weeks. Pressian. 2021 October 5.

17. Ko KK, W. HJ, M. PJ. Comparative Study of the Trend and Policy: Response of COVID-19 in Global Cities. Korean Association for Local Government Studies. 2021;33(2):93–118.

18. Gerritse M. COVID-19 transmission in cities. Eur Econ Rev. 2022;150:104283.

19. Tucker P, Gilliland J. The effect of season and weather on physical activity: a systematic review. Public Health. 2007;121(12):909–22.

20. Shin JH. From June 1st, phase 1 of ‘support for daily recovery of vaccinated people. Korea Policy Briefing. 2021 May 31.

21. Snoeijer B, Burger M, Sun S, Dobson R, Folarin A. Measuring the effect of Non-Pharmaceutical Interventions (NPIs) on mobility during the COVID-19 pandemic using global mobility data. npj Digital Medicine. 2021;4.

22. Kim KT, Lee IM, Kwak HC, Min JH. Application study of telecommunication record data in floating population estimation. Seoul Studies. 2015;16(3):177–87.

23. Korea S. Overall economically active population by gender. Statistics Korea2023.

24. Holding a meeting of representative organizations by industry to spread telecommuting [press release]. Ministry of Employment and Labor, December 23 2021.

